# S-MiXcan: Inferring Cell-Type-Level Transcriptome-Wide Associations from Bulk Transcriptomics Using GWAS Summary Statistics

**DOI:** 10.64898/2026.03.22.26349035

**Authors:** Sinan Zhu, Qiao Fan, Xiaoyu Song

**Affiliations:** Centre for Biomedical Data Science, Duke-NUS Medical School, National University of Singapore

**Keywords:** cell type, transcriptome-wide association study, summary statistics, association probability

## Abstract

Cell-type–specific regulation of gene expression plays a central role in complex disease etiology, yet most transcriptome-wide association studies (TWAS) rely on bulk tissue models. Recently, a couple methods leverage single-cell transcriptomics to perform TWAS at the cell-type resolution, but they are limited by scarce matched genotype– single-cell cohorts and restricted to peripheral blood, with minimal coverage of less accessible, disease-relevant tissues. In this study, we developed S-MiXcan, a summary-statistics–based TWAS framework that enables cell-type–aware association analysis using bulk transcriptomic data across *K* ≥ 2 cell types without requiring individual-level data. As a major advancement over our prior tool MiXcan, S-MiXcan jointly models genetically regulated expression (GReX) across *K* cell types, accounts for cross–cell-type correlations, identifies disease-associated genes, and provides probabilistic interpretations for distinguishing cell-type–specific from shared associations. In real data analyses, compared with using individual-level genotype-based implementation, S-MiXcan achieved highly concordant results (Pearson’s *r* ≈ 1) in cell-type-aware TWAS. Applied to large-scale multi-cohort Genome-Wide Association Study (GWAS) meta-analyses from the Breast Cancer Association Consortium, S-MiXcan maintained well-controlled type I error (genomic inflation *λ* = 1.057), identified key breast cancer risk associated genes that function in a cell-type specific manner, and revealed relevant cell types through probabilistic inference. These results demonstrate that S-MiXcan, publicly available at https://github.com/songxiaoyu/SMiXcan, provides a scalable and interpretable framework for cell-type–aware TWAS.

---

Transcriptome-wide association studies (TWAS) provide a powerful two-sample framework for identifying disease-associated genes, by training genetically regulated expression (GReX) prediction models in one cohort and testing disease associations using predicted GReX levels in another independent cohort. By aggregating signals from millions of genetic variants into gene-level associations, TWAS substantially alleviates the multiple-testing burden inherent to genome-wide association studies (GWAS), thereby increasing statistical power, facilitating biological interpretation, and highlighting signals in the disease relevant tissues. Nevertheless, conventional TWAS methods, such as PrediX-can [1] and FUSION [2], perform analyses at the bulk tissue level, treating tissues as homogeneous and ignoring underlying cell-type heterogeneity. This simplification can compromise the accuracy of GReX prediction and obscure true disease associations, particularly when disease-relevant cell types constitute only a small fraction of the tissue [3].

Recent methodological advances have begun to address these limitations by modeling genetic effects at the cell-type level. cWAS [4] infers cell-type proportions from bulk expression and links genetically regulated proportion changes to disease; this approach, however, does not investigate disease-associated genes or gene-level regulatory effects. MiXcan [3] similarly decomposes bulk expression into cell-type–specific components but performs cell-type-aware TWAS to pinpoint trait-associated genes and their functional cell types; nonetheless, it is limited to two cell types (the cell type of interest versus all others) and requires access to individual-level genotype datasets. In contrast, scTWAS [5] performs TWAS directly at single-cell resolution by leveraging single-cell transcriptomic data, and scPrediXcan [6] further incorporates functional genomic annotations to improve expression prediction in single cells. Both single-cell-based approaches, however, require matched genotype and single-cell transcriptomics datasets in a sufficiently large number of individuals and are currently applicable primarily to peripheral blood, with limited coverage for rarer tissues in existing atlases. For example, within the Genotype-Tissue Expression (GTEx) consortium, matched genotype and snRNA-seq data for normal mammary tissue are available only for three individuals [7].

In this study, we introduce S-MiXcan, a major extension of MiXcan for a summary-statistics-based cell-type-aware TWAS using bulk transcriptomics data. It exhibits three major advancements over MiXcan. First, it enables summary statistics-based inference by explicitly accounting for correlations among estimated cell-type–specific GReX weights, which arise from cell-type decomposition uncertainty, finite-sample estimation variability, and shared genomic predictors across cell types, and thereby ensuring valid and well-powered association tests. Second, S-MiXcan substantially improves the interpretability of cell-type specific versus shared associations by quantifying association probabilities for different cell-type patterns, providing informative measures of cell-type specificity that go beyond p-values. Lately, it also systematically updates the modeling and inference strategies, such as supporting joint estimation and inference across three or more cell types. Together, these features enable S-MiXcan to perform highly confident cell-type-aware TWAS while preserving data privacy, improving computational efficiency and robustness, enhancing interpretability of the discoveries, and facilitating large-scale meta-analyses.

## Schematic overview of S-MiXcan

S-MiXcan operates in two stages (Figure 1). In the first stage, S-MiXcan uses a training dataset that comprises of matched genotype and bulk transcriptome data to train cell-type-level GReX prediction models. It decomposes the observed gene expression levels into multiple cell types, estimates cell-type proportions, and learns cell-type-level GReX prediction models for each gene jointly across all cell types. In the second stage, S-MiXcan leverages GWAS summary statistics from an independent cohort, as well as the linkage disequilibrium (LD) structure from a reference genome, to infer disease associations with each GReX levels in different cell types. It accounts for cell-type correlations, derives tissue-level gene–disease association p-values, and infers post association probabilities for each cell type pattern.

**Figure 1:**
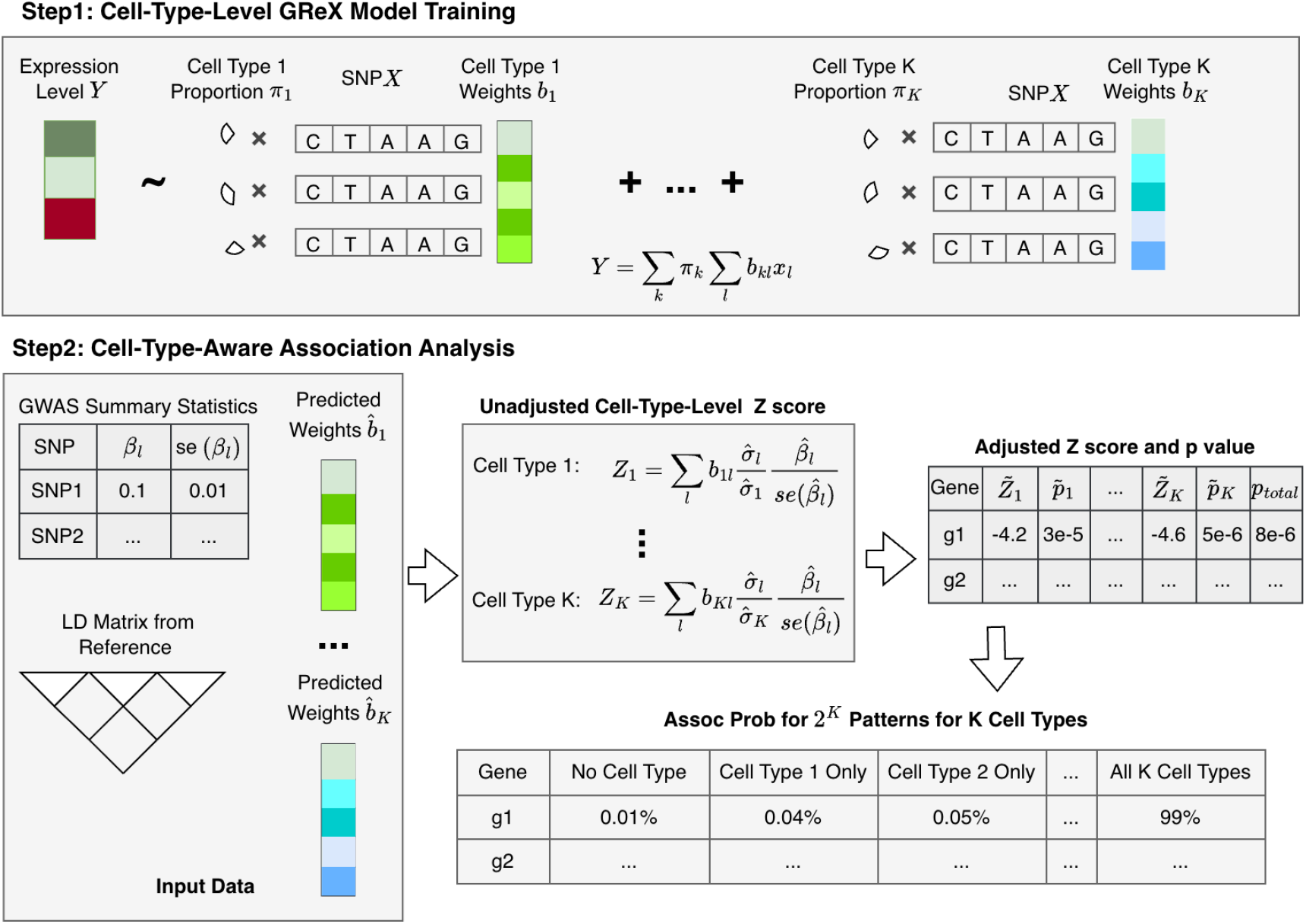
Schematic overview of S-MiXcan. S-MiXcan trains cell-type-level GReX prediction models in a matched genotype-expression dataset, and applies the cell-type-level training weights to an independent GWAS summary statistics to identify trait-associated genes and infer their functional cell types.

## Cell-type-level GReX prediction model

Extending MiXcan, we develop cell-type-level GReX prediction models for *K* ≥ 2 cell types. Suppose we have *n* bulk tissue samples composed of a mixture of *K* cell types. For each sample *i* = 1, …, *n*, let *π*_*ik*_ denote the proportion of cell type *k* ∈ {1, …, *K*} in the sample *i*, where 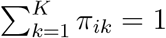. Let *y*_*i*_ denote the standardized bulk expression level of a gene, and let **x**_*i*_ = (*x*_*i*1_, …, *x*_*iP*_)^⊤^ ∈ ℝ^*P*^ denote the vector of genotypes within the pre-specified neighborhood of the gene, such as the single nucleotide polymorphisms (SNPs) within 1MB of the transcription start site of the gene. Without loss of generalizability, we omit covariates, which can be added as additional predictors in regressions. With cell-type decomposition, the bulk expression *y*_*i*_ can be written as a weighted average of gene expression in each cell type, such that 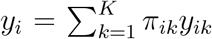, where *y*_*ik*_ denotes the unobserved expression for sample *i* in cell type *k*.

For each cell type, we assume a linear genetic model 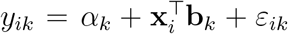, where *α*_*k*_ ∈ ℝ is a cell-type–specific intercept, **b**_*k*_ ∈ ℝ^*P*^ is the cell-type–specific genetic effects on expression, and *ε*_*ik*_ is a mean-zero error term. Linear model is considered here not only for its simplicity, but also because it has been shown to outperform deep learning, random forest, and other complex machine learning methods in this setting [8]. Plugging the cell-type-level linear genetic model into 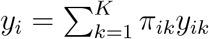, we have

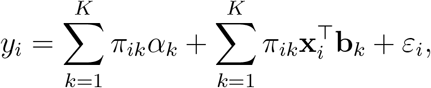

where 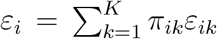 is the model error. This equation has two sets of parameters *π*_*ik*_ and (*α*_*k*_, **b**_*k*_). The parameter *π*_*ik*_ is a tissue-level parameter that’s shared across all genes, and thus we estimate jointly across genes via deconvolution. Relaxed from MiXcan, S-MiXcan can be integrated with any existing transcriptomics-based decomposition strategy. For example, we use BayesDeBulk [9] for its capacity to model *K* ≥ 2 cell types with robustness against misspecificaition of expression profiles.

With estimated 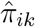, we next estimate the gene-specific parameters (*α*_*k*_, **b**_*k*_) by adopting a mean-plus-contrast reparameterization strategy. Specifically, let **C** ∈ ℝ^*K*×(*K*−1)^ be a full-column-rank contrast matrix satisfying **C**^⊤^**1**_*K*_ = **0** (e.g., Helmert contrasts). Let 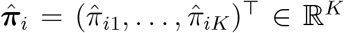 be the estimated cell-type proportion for sample *i*, then its contrast-coded composition ***ĉ***_*i*_ is a vector of length *K* − 1 such that 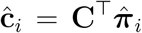. The transformation of ***π***_*i*_ to ***ĉ***_*i*_ ensures symmetry in the penalized regression and identifiability under the compositional constraint. With the reparameterization, we can write 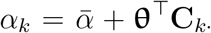, where **C**_*k*·_ ∈ ℝ^*K*−1^ is defined as the *k*-th row of 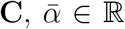 represents the average intercept, and**θ** ∈ ℝ^*K*−1^ captures the cell-type deviations from this intercept. Similarly, we can write 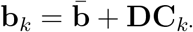, where 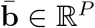 represents the average genetic effects and **D** ∈ ℝ^*P* ×(*K*−1)^ characterizes the cell-type–specific deviations from these average effects.

Substituting these parameters and reorganizing the predictors, we can obtain the working regression

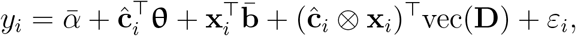

where ⊗ denotes the Kronecker product and vec(**D**) ∈ ℝ^*P* (*K*−1)^ stacks the columns of **D** ∈ ℝ^*P* ×(*K*−1)^. The new parameters 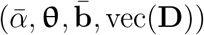 are jointly estimable via elastic-net regression, where we put penalties on 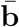 and vec(**D**) to penalize genetic effects on expression to improve their prediction performance. Finally, with the estimated parameters, we can construct the cell-type–specific GReX weights as 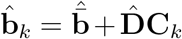 for each cell type *k* = 1, …, *K*. Genes with at least one 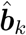 nonzero across the *K* cell types are regarded as genetically predictable and assessed for their association with the phenotypes in the downstream analyses.

## GWAS summary-based cell-type-level association before cell-type correlation adjustment

GWAS summary statistics–based inference is well established in tissue-level TWAS [1]. However, prior work has shown that naively extending tissue-level TWAS strategies to cell-type–specific analyses, without accounting for correlations among cell types, can lead to inflated type I error [3]. To address this, we propose a two-step framework. First, we derive GWAS summary statistics–based association tests for cell-type–level GReX by extending existing TWAS methodology while temporarily ignoring cross–cell-type correlations. Second, we develop a correlation-adjustment procedure that explicitly models dependencies among cell types to ensure valid inference.

Specifically, suppose we have GWAS summary statistics results derived from an independent cohort, which includes z-score for each genotype that capture both the association directions and significance. For a given gene, consider the GWAS phenotype 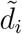 and genotypes 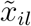 for SNP *l* = 1, …, *P* in the pre-defined neighborhood of the gene. For cell type *k*, the predicted GReX is 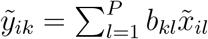, where *b*_*kl*_ is cell-type–specific GreX weights estimated from training data. Our goal is to identify phenotype-associated genes and their functional cell types, by by leveraging these pre-trained cell-type–specific GReX prediction models.

Without loss of generalizability, we consider a marginal linear model, where only one cell type is modeled at a time, and we have

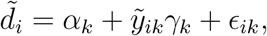

whose corresponding Z-score, 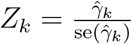, captures disease association in cell type *k*.

In the absence of individual-level genotypes, we derive a Z-score from GWAS summary statistics by borrowing the idea of S-PrediXcan [1](see Supplementary Materials for details), and we obtain

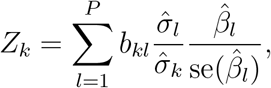

where *b*_*kl*_ is the predicted weight of SNP *l* in cell type *k* for the gene from the training model; 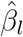 and 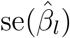 are the estimate and standard error of the SNP *l* in the GWAS summary statistics; 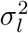 is variance of SNP *l* estimable from a reference genome; and finally 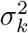 is the variance of predicted GReX for cell type *k*. To estimate 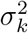, we let **b**_*k*_ be the the vector of SNP weights for cell type *k* and **Γ**_*g*_ be the variance-covariance matrix of these SNPs estimated from the LD matrix of the reference genome, and then the variance of predicted GReX can be estimated such that 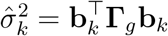.

## Correlation-adjusted cell-type–level association

The marginal association statistic *Z*_*k*_ is obtained by testing each cell type separately. However, because predicted GReX across cell types are constructed from overlapping genetic predictors and inferred from the same bulk expression data, the resulting marginal Z-scores (*Z*_1_, …, *Z*_*K*_) are generally correlated [3], reflecting both biological co-regulation and technical dependencies from decomposition uncertainty in finite samples.

To account for this correlation, we consider a joint linear model that simultaneously models disease associations across all *K* cell types, such that

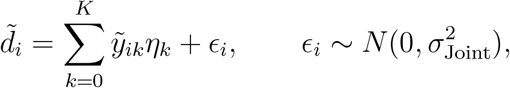

where 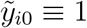 denotes the intercept, and **η** = (*η*_0_, *η*_1_, …, *η*_*K*_)^⊤^ is the vector of regression coefficients corresponding to the intercept and slopes in *K* cell types. Without individual genotype data, our goal is to use the original marginal Z-scores **Z** = (*Z*_1_, …, *Z*_*K*_)^⊤^, where each 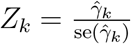 in the marginal model, to estimate the joint Z-scores 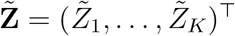, where each 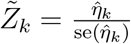 in the joint model.

We assume that each gene explains only a small proportion of phenotypic variance, as is commonly observed in complex human diseases and generally assumed in TWAS methods such as PrediXcan; statistically, this corresponds to a small coefficient of determination (*R*^2^). Then, the joint cell-type–specific association statistics 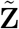 can be well approximated from a linear transformation of the marginal Z-scores **Z** (Derivation in Supplementary Materials) such that

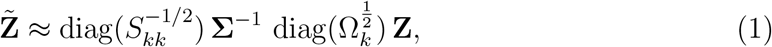

where 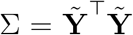 is the variance-covariance matrix of the *n* × (*K* + 1) design matrix 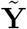 including ones as the first column and GReX as the rest *K* columns; 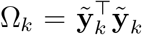 is the variance of 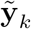, the GReX value in cell type *k*; and *S*_*kk*_ is the *k*-th diagonal element of Σ^−1^, ensuring that each 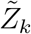 is properly standardized.

Please note that predicted GReX across cell types may be highly correlated, causing collinearity problem in the equation, and we apply ridge regularization to stabilize estimation such that 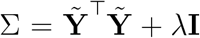 and 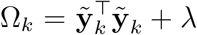, where *λ >* 0 is a small adaptive regularization parameter that increases shrinkage under stronger collinearity.

## Determination of significance at the tissue level

Cell-type–specific two-sided p-values are computed from the correlation-adjusted Z-scores as

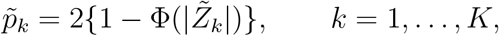

where Φ(·) denotes the standard normal cumulative distribution function.

To summarize evidence of association across cell types while allowing for arbitrary dependence among test statistics, we apply the aggregated Cauchy association test (ACAT) to give tissue-level p-value. Specifically, given the cell-type–specific p-values 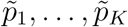, the ACAT statistic is defined as

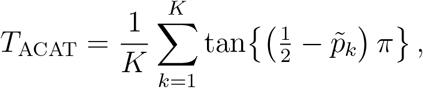

and the corresponding tissue-level p-value for this gene is computed as

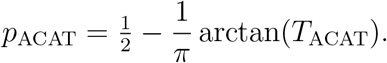

The resulting ACAT p-value serves as a composite test for gene–disease association in any cell types in this tissue, which can be further adjusted for multiple testing when multiple genes are simultaneously evaluated for transcriptome-wide associations. It is worth noting that while we develop an adjustment strategy for correlations across cell types, the cell-type-level p-values can still be inflated under leakage of non-linear signals from other cells that could not be fully adjusted. Therefore, tissue-level p-value is recommended to conclude the gene–disease association.

## Inferring cell-type–specific association patterns

While the ACAT p-value indicates whether a gene is associated with disease in any cell type within a tissue, it does not clarify whether the association is cell-type–specific or shared. To facilitate such interpretation post tissue-level inference, we applied the probabilistic pattern-based framework [11], which accounts for cross cell-type correlations to estimate association probabilities.

Specifically, for *K* cell types, we define *L* = 2^*K*^ mutually exclusive binary association patterns **q**_*ℓ*_ ∈ {0, 1}^*K*^, where *q*_*ℓk*_ = 1 indicates association in cell type *k* and 0 otherwise. For example, when *K* = 2, we have *L* = 4 and the four patterns are 00 (no association), 10 (cell type 1 only), 01 (cell type 2 only), and 11 (shared association). For each gene, we transform the *K* cell-type–specific p-values as *t*_*k*_ = −2 log(*pk*). Under the null, 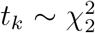, whereas under the alternative it follows a mixture of non-central chi-squared distribution, estimable using a scaled chi-squared distribution in Primo [11]. We then estimate the null and alternative density functions, *f*_0*k*_ and *f*_1*k*_, for each cell type *k*, and use them to construct pattern-specific densities and compute the association probabilities that each gene belongs to each of the *L* association patterns (see Supplementary Materials for details).

## Model training in GTEx mammary tissue samples

Next, we trained GReX prediction models using S-MiXcan based on 125 mammary tissue samples from women of European ancestry (EA) in the GTEx consortium v8 [12]. As a proof of concept, we restricted our analyses to 6,443 genes and their neighboring genetic variants that were previously examined [3], where data processing procedures followed the original publications[12, 3]. In model training, we first decomposed the mammary tissue into epithelial and stromal compartments, as epithelial cells are the primary origin of breast cancer while stromal cells provide structural and microenvironmental support. We curated epithelial marker genes to estimate epithelial proportion and major stromal components, such as adipocyte, fibroblast and endothelial marker genes, to estimate stromal proportion (Supplementary Table 1).

We then applied BayesDeBulk [9] to infer cell-type proportions from bulk expression data. With the estimated proportions, we jointly model the cell-type–level genetic effects on gene expression with 10-fold cross-validation, controlling for covariates including age, sequencing platform, PCR batch, genomic principal components (PCs) 1–5, and the top 15 PEER factors. The models yielded GReX prediction models for 6,405 of the 6,443 genes, each having at least one non-zero SNP weight in at least one cell type. Of these, 4,734 exhibited cell-type-specific genetic regulation (distinct weights across the two cell types), whereas 1,671 showed shared regulation. These 6,405 GReX models were subsequently leveraged to identify breast cancer risk–associated genes.

## Comparison with individual-level genotypes

To demonstrate that S-MiXcan can produce similar results comparable to those obtained using individual-level genotype data as in MiXcan, which exhibited well-controlled Type I error and unique identifications [3], we next performed both MiXcan and S-MiXcan association analyses in the Discovery, Biology, and Risk of Inherited Variants in Breast Cancer (DRIVE) cohort [13]. The cohort included genotype data profiled by OncoArray-30K for 58,648 women with European ancestry (EA) including 31,716 cases and 26,932 controls. To apply S-MiXcan, we first calculated the GWAS summary statistics based on the individual-level genotype data, then applied the mammary tissue GTEx GReX prediction models to the DRIVE GWAS summary statistics referencing its own LD matrix. For comparison, we also applied the GReX prediction models directly to the individual-genotype data using MiXcan. The results of MiXcan and S-MiXcan demonstrated a high concordance (Figure 2), with the Pearson correlation of -log10(p-value) > 0.99, demonstrating that S-MiXcan can robustly produce cell-type-aware TWAS results in absence of individual-level genotype data.

**Figure 2:**
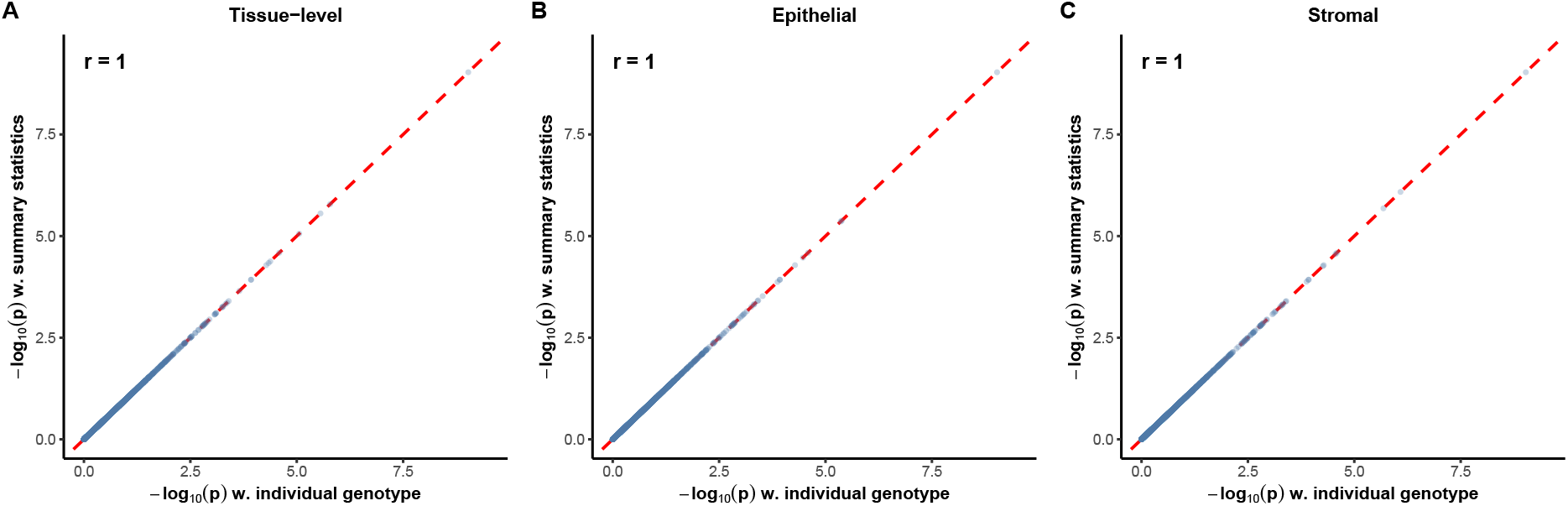
S-MiXcan based on summary statistics produces p-values highly concordant with those obtained from individual-level genotype data in the DRIVE cohort. (A) Scatter plot comparing *log*_10_(*p*) obtained from individual-level genotype data in MiXcan versus summary statistics in S-MiXcan across at the tissue level. Each point represents one gene, and the dashed red line indicates the line of equality (y = x). The Pearson correlation coefficient (r) quantifies concordance between the two methods. (B) Scatter plot comparing *log*_10_(*p*) obtained from individual-level genotype data in MiXcan versus summary statistics in S-MiXcan in epithelial cells. (C) Scatter plot comparing *log*_10_(*p*) obtained from individual-level genotype data in MiXcan versus summary statistics in S-MiXcan in stromal cells.

## Cell-type-aware TWAS for breast risk using GWAS summary statistics from Breast Cancer Association Consortium (BCAC)

To demonstrate the utility of S-MiXcan, we next applied it to the GWAS summary statistics from the large-scale BCAC meta-analysis of breast-cancer risk in EA women [14], which comprises 228,951 participants (122,977 cases and 105,974 controls) with SNPs matching the 1000 Genomes Project Phase 3 [15]. LD matrices were constructed using the EA reference panel from the same project. The resulting genomic inflation factor was estimated as 1.058 (Figure 3A) using the Bayesian method BACON [16], indicating a well-controlled type I error. Out of 6,405 genes with non-zero prediction weights, S-MiXcan identified 32 genome-wide significant genes at a 5% family-wise error rate (FWER; *p <* 7.8 × 10^−6^) and 76 suggestive genes at 10% false discovery rate (FDR; *p <* 8.3 × 10^−4^) (Supplementary Table 2). The association probabilities for these 76 suggestive genes suggested 71 of them presented non-identical effects in two cell types (Figure 3B), including one gene (SRGAP1) pre-dominatingly in epithelial cells, seven genes (FES, CTSW, EP300, TMEM69, CENPS, FKBP2, IQCH-AS1) in stromal cells, and the rest 63 genes with different estimated effects but function in both cells (Figure 3B). Among them, FES, CTSW, and EP300 have >95% probabilities of only acting in stromal cells.

**Figure 3:**
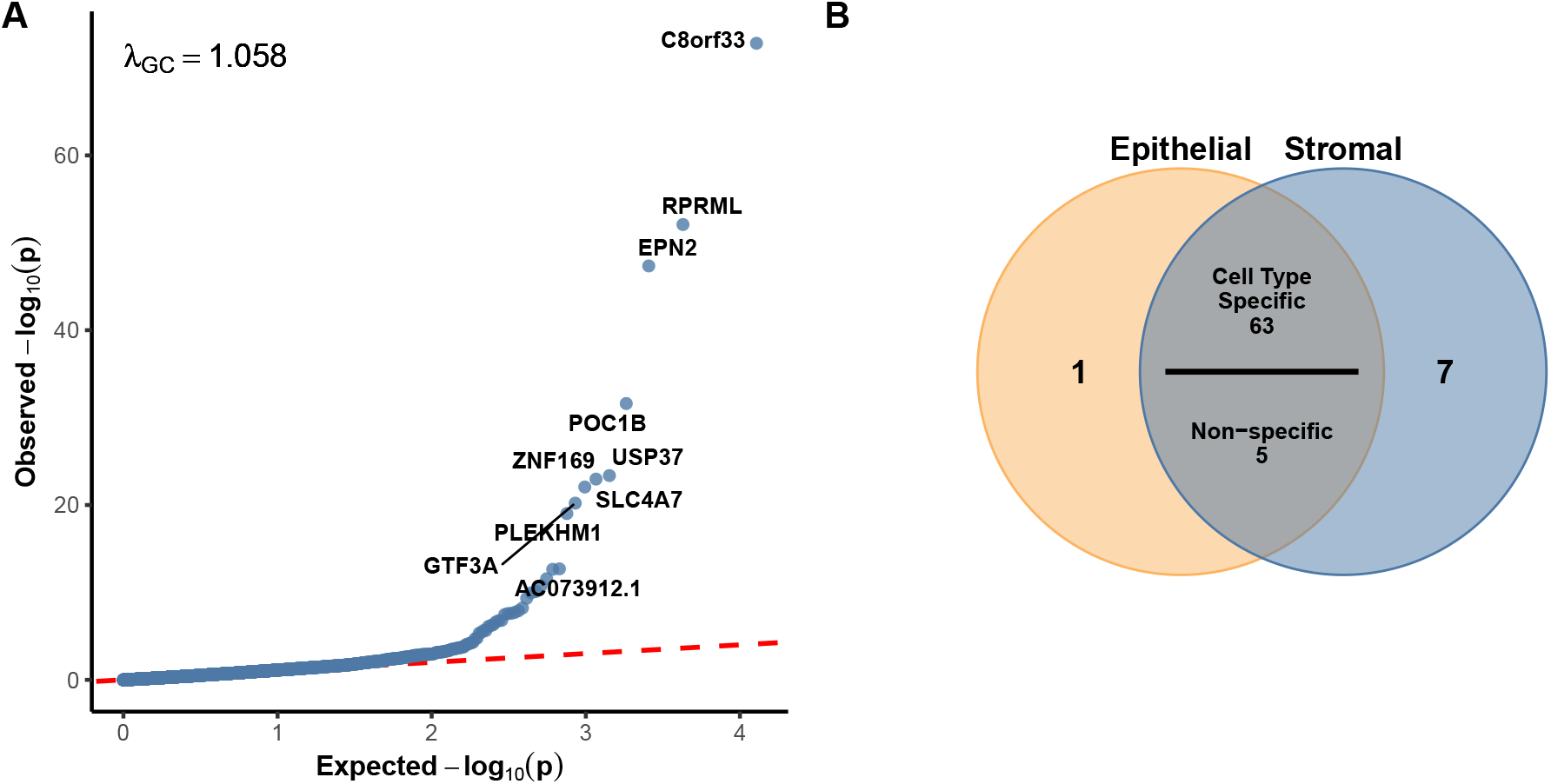
S-MiXcan demonstrates well controlled Type I error and cell-type-specific gene discovery. (A) Application of S-MiXcan to BCAC meta-analysis GWAS summary statistics maintains well-controlled type I error (*λ*_*GC*_ = 1.058) with significant hits. (B) Inferred cell-type-specificity among 76 suggestive genes at 10% FDR.

FES is an oncogenic tyrosine kinase that promotes angiogenesis and macrophage-driven metastasis in mouse models [17], and we found its stormal expression positively associated with breast cancer risk. CTSW is an immune cell–specific tumor suppressor linked to improved breast cancer survival [18], and we observed its stormal expression was negatively associated with risk. EP300, a histone acetyltransferase with context-dependent tumor suppressor and oncogenic roles [19], showed a negative association with breast cancer risk when expressed in stromal cells.

## Cell-type-aware TWAS performance under *K >* 2 cell types

Although S-MiXcan can in principle be applied to *K* cell types, its statistical power may decline in small samples when the number of cell types is large. This is because the number of parameters in the regression models increases substantially with the number of cell types and the cell-type decomposition becomes more complex to estimate, leading to greater estimation uncertainty and amplified multicollinearity across predicted GReX. To illustrate this, we further decomposed the 125 mammary tissues into three cell types, including epithelial cells, adipocytes/endothelialc cells, and fibroblasts, estimated their proportions, constructed cell-type–specific prediction models, and applied these models to the DRIVE study with individual-level genotype data as well as to BCAC breast cancer risk GWAS summary statistics. In total, 6,403 genes had non-zero prediction weights, similar to the two–cell-type models. The predicted GReX from summary-statistics–based models remained highly correlated with those derived from individual-level genotype data at the tissue and cell type levels (all − *log*_10_(*p*) r>0.99; Figure S1). The genomic inflation factor was 1.064 (Figure S2A), indicating well-controlled type I error. However, the three cell-type S-MiXcan identified 17 genome-wide significant genes at a 5% FWER (p < 7.8 × 10−6) and 33 suggestive genes at 10% FDR (p < 4 × 10−4) (Supplementary Table 3; Figure S2A), demonstrating a substantial loss of power relative to the two–cell-type analysis. Cell type specificity remained for the three cell type models (Figure S2B).

## Discussion

We developed S-MiXcan, a summary-statistics-based TWAS method that enables cell-type-aware analysis across *K* ≥ 2 cell types without requiring individual-level genotype data and provides probabilistic inference of cell-type–specificity. Compared with MiXcan, S-MiXcan introduces three key advances: (1) scalability to large multi-cohort GWAS meta-analyses using summary statistics; (2) extension to *K >* 2 cell types through revised cell-type composition estimation and cell-type–level prediction modeling; and (3) improved interpretability via a probabilistic framework for characterizing cell-type–specific versus shared associations. Real data based evaluation demonstrated that S-MiXcan achieves comparable performance to individual-level genotype-based analyses, maintains well-controlled type I error, identifies key risk genes, and infers associated cell types with high confidence. Potential limitations include prediction variations with small training samples relative to the number of cell types. Future work could incorporate larger and more diverse cohorts, and integrate additional omics data to further enhance robustness and biological resolution.

## Data Availability

S-MiXcan software, documentation, and tutorial are open-source at https://github.com/songxiaoyu/SMiXcan. The data used in this study are publicly available. GTEx data are available from GTEx v8 (dbGaP accession number phs000424.v8.p2 (https://www.ncbi.nlm.nih.gov/projects/gap/cgi-bin/study.cgi?study_id=phs000424.v8.p2). Discovery, Biology, and Risk of Inherited Variants in Breast Cancer (DRIVE) data are available from dbGaP accession number phs001265.v1.p1 (https://www.ncbi.nlm.nih.gov/projects/gap/cgi-bin/study.cgi?study_id=phs001265.v1.p1). Breast cancer risk GWAS summary statistics are available from https://www.ccge.medschl.cam.ac.uk/breast-cancer-association-consortium-bcac/data-data-access/summary-results/gwas-summary-associations. The 1000 Genomes Project Phase 3 reference data are available at https://www.internationalgenome.org/cate.

https://www.ccge.medschl.cam.ac.uk/breast-cancer-association-consortium-bcac/data-data-access/summary-results/gwas-summary-associations

https://www.internationalgenome.org/cate

https://github.com/songxiaoyu/SMiXcan

## Code and Data Availability

S-MiXcan software, documentation, and tutorial are open-source at https://github.com/songxiaoyu/SMiXcan. GTEx data are available from GTEx v8 (dbGaP accession number phs000424.v8.p2 (https://www.ncbi.nlm.nih.gov/projects/gap/cgi-bin/study.cgi?study_id=phs000424.v8.p2). Discovery, Biology, and Risk of Inherited Variants in Breast Cancer (DRIVE) data are available from dbGaP accession number phs001265.v1.p1 (https://www.ncbi.nlm.nih.gov/projects/gap/cgi-bin/study.cgi?study_id=phs001265.v1.p1). Breast cancer risk GWAS summary statistics are available from https://www.ccge.medschl.cam.ac.uk/breast-cancer-association-consortium-bcac/data-data-access/summary-results/gwas-summary-associations. The 1000 Genomes Project Phase 3 reference data are available at https://www.internationalgenome.org/cate.

## Acknowledgement

This work was supported by Singapore Ministry of Education FY2024-MOET1-0004, Singapore Ministry of Health Duke-NUS Signature Research Programme, and Singapore National Medical Research Council (NMRC) CS-IRG CIRG25jul-0023.

