## Supplementary Material for "S-MiXcan: Inferring Cell-Type-Level Transcriptome-Wide Associations from Bulk Transcriptomics Using GWAS Summary Statistics"

#### S1. Technical details for "GWAS summary-based cell-type-level association before cell-type correlation adjustment".

This section provides technical details for leveraging cell-type-level genetically regulated expression (GReX) prediction models trained in a matched genotype–expression dataset to test for disease associations using GWAS summary statistics derived from an independent cohort, prior to adjusting for correlations across cell types. The impact of cell-type correlations is derived in subsequent sections.

Consider an independent GWAS cohort with $n$ individuals indexed by $i = 1, \dots, n$. Let $\tilde{d}_i \in \mathbb{R}$ denote the phenotype value for individual $i$, and let $\tilde{x}_{il} \in \mathbb{R}$ denote the genotype dosage for individual $i$ at SNP $l$, where $l = 1, \dots, P$, within the cis-region of the gene. Let $K$ denote the number of cell types under consideration. For a given cell type $k \in \{1, \dots, K\}$, the predicted GReX level for individual $i$ is defined as

$$\tilde{y}_{ik} = \sum_{l=1}^{P} b_{kl}\, \tilde{x}_{il}, \tag{1}$$

where $b_{kl}$ denotes the weight for SNP $l$ on the gene in cell type $k$, estimated from the training dataset.

Ignoring correlations across cell types, we model the association between the phenotype and predicted GReX for each cell type $k$ using a marginal linear model as follows:

$$\tilde{d}_i = \tilde{y}_{ik}\gamma_k + \epsilon_{ik}, \qquad \epsilon_{ik} \sim N(0, \sigma^2_{\epsilon_k}), \tag{2}$$

where $\gamma_k$ denotes the marginal association effect of predicted GReX in cell type $k$, and $\sigma^2_{\epsilon_k}$ denotes the residual variance in the marginal model.

Although individual-level genotypes are unavailable and we are unable to directly impute $\tilde{y}_{ik}$, we can derive the marginal association Z-score for each cell type $k$ from the GWAS summary statistics defined as

$$Z_k = \frac{\hat{\gamma}_k}{\text{se}(\hat{\gamma}_k)}. \tag{3}$$

To obtain $Z_k$, we first derive the numerator $\hat{\gamma}_k$ and the denominator $\text{se}(\hat{\gamma}_k)$ separately and then calculate their ratio.

#### Derivation of $\hat{\gamma}_k$

Under ordinary least squares, $\hat{\gamma}_k$ can be obtained as follows

$$\hat{\gamma}_k = \frac{\sum_{i=1}^{n} \tilde{y}_{ik}\tilde{d}_i}{\sum_{i=1}^{n} \tilde{y}_{ik}^2}. \tag{4}$$

Substituting $\tilde{y}_{ik} = \sum_{l=1}^{P} b_{kl}\tilde{x}_{il}$, the numerator of Equation (4) can be rewritten as

$$\text{Numerator} = \sum_{i=1}^{n}(\sum_{l=1}^{P} b_{kl}\tilde{x}_{il}\tilde{d}_i) = \sum_{l=1}^{P} b_{kl}\sum_{i=1}^{n}\tilde{x}_{il}\tilde{d}_i. \tag{5}$$

To estimate the $\sum_{i=1}^{n}\tilde{x}_{il}\tilde{d}_i$ component of the numerator, we look in to the GWAS summary statistics. Let $\hat{\beta}_l$ denote the marginal SNP effect estimate, $\text{se}(\hat{\beta}_l)$ its standard error, and $\hat{\sigma}_l^2$ the variance of SNP $l$, estimated from an external reference panel. Then, similarly with ordinary least squares, we have

$$\hat{\beta}_l = \frac{\sum_{i=1}^{n}\tilde{x}_{il}\tilde{d}_i}{n\hat{\sigma}_l^2}. \tag{6}$$

Therefore,

$$\sum_{i=1}^{n}\tilde{x}_{il}\tilde{d}_i = n\hat{\sigma}_l^2\hat{\beta}_l. \tag{7}$$

Plugging Equation (7) in to the Equation (4) and (5), we have

$$\hat{\gamma}_k = \frac{\sum_{l=1}^{P} b_{kl}\hat{\beta}_l\hat{\sigma}_l^2}{\hat{\sigma}_k^2}, \tag{8}$$

where $\hat{\sigma}_k^2$ is the estimated variance of predicted GReX in cell type $k$ is $\hat{\sigma}_k^2 = \frac{1}{n}\sum_{i=1}^{n}\tilde{y}_{ik}^2$.

#### Derivation of $\text{se}(\hat{\gamma}_k)$

Under the marginal linear model, our goal is to link $\text{se}(\hat{\gamma}_k)$ with GWAS summary statistics, where $\text{se}^2(\hat{\gamma}_k)$ is defined such that

$$\text{se}^2(\hat{\gamma}_k) = \frac{\sigma_{\epsilon_k}^2}{n\hat{\sigma}_k^2}. \tag{9}$$

To estimate $\sigma_{\epsilon_k}^2$, we first introduce $\hat{\sigma}_d^2$ denoting the total phenotypic variance. Given the marginal linear model for each cell type $k$, we can obtain $R_k^2$, the the proportion of the total variation $\hat{\sigma}_d^2$ explained by the gene expression in the single cell type $k$, as follows

$$R_k^2 = \hat{\gamma}_k^2\frac{\hat{\sigma}_k^2}{\hat{\sigma}_d^2}, \tag{10}$$

where $\gamma_k$ is the estimated marginal regression coefficient of the gene in cell type $k$ on the phenotype, $\hat{\sigma}_k^2$ is the sample variance of the gene in cell type $k$. Then, the remaining variance $\sigma_{\epsilon_k}^2$, the proportion of the total variation $\hat{\sigma}_d^2$ not explained the gene expression in cell type $k$, can be written as follows:

$$\sigma_{\epsilon_k}^2 = \hat{\sigma}_d^2(1 - R_k^2). \tag{11}$$

Plugging Equation 11 into 9, we have

$$\mathrm{se}^2(\hat{\beta}_l) = \frac{\sigma^2_{\epsilon_k}}{n\hat{\sigma}^2_k} = \frac{\hat{\sigma}^2_d(1-R^2_k)}{n\hat{\sigma}^2_k}. \tag{12}$$

Similarly, leveraging GWAS summary statistics, we can obtain $R^2_l$, the the proportion of the total variation $\hat{\sigma}^2_d$ explained by a single SNP $l$, as follows

$$R^2_l = \hat{\beta}^2_l \frac{\hat{\sigma}^2_l}{\hat{\sigma}^2_d}, \tag{13}$$

where $\beta_l$ is the estimated marginal regression coefficient of SNP $l$ on the phenotype, $\hat{\sigma}^2_l$ is the sample variance of the genotype at SNP $l$. Following the same strategy, we have

$$\mathrm{se}^2(\hat{\beta}_l) = \frac{\hat{\sigma}^2_d(1-R^2_l)}{n\hat{\sigma}^2_l}. \tag{14}$$

Leverage Equation (14) to replace $\hat{\sigma}^2_d$ from Equation (12), we have

$$\mathrm{se}^2(\hat{\gamma}_k) = \frac{(1-R^2_k)\hat{\sigma}^2_l\,\mathrm{se}^2(\hat{\beta}_l)}{(1-R^2_l)\hat{\sigma}^2_k}. \tag{15}$$

#### Final marginal Z-score

Putting $\hat{\gamma}_k$ and $\mathrm{se}(\hat{\gamma}_k)$ together, $Z_k = \frac{\hat{\gamma}_k}{\mathrm{se}(\hat{\gamma}_k)}$ can be written as follows:

$$Z_k = \sum_{l=1}^{P} b_{kl} \frac{\hat{\sigma}_l}{\hat{\sigma}_k} \frac{\hat{\beta}_l}{\mathrm{se}(\hat{\beta}_l)} \sqrt{\frac{1-R^2_l}{1-R^2_k}}. \tag{16}$$

Under complex human diseases, both $R^2_l$ and $R^2_k$ are typically small, we have the approximation of z-score for cell type $k$ such that

$$Z_k \approx \sum_{l=1}^{P} b_{kl} \frac{\hat{\sigma}_l}{\hat{\sigma}_k} \frac{\hat{\beta}_l}{\mathrm{se}(\hat{\beta}_l)}, \tag{17}$$

where $b_{kl}$ is the predicted weight of SNP $l$ in cell type $k$ for the gene from the training model; $\hat{\beta}_l$ and $\mathrm{se}(\hat{\beta}_l)$ are the estimate and standard error of the SNP $l$ in the GWAS summary statistics; $\sigma^2_l$ is variance of SNP $l$ estimable from a reference genome; and finally $\sigma^2_k$ is the variance of predicted GReX for cell type $k$.

To estimate $\hat{\sigma}^2_k$, we let $\mathbf{b}_k = (b_{k1}, \ldots, b_{kP})^\top \in \mathbb{R}^P$ denote the weight vector for cell type $k$, and let $\mathbf{\Gamma}_g \in \mathbb{R}^{P\times P}$ denote the linkage disequilibrium covariance matrix of the SNPs estimated from a reference panel. Then

$$\hat{\sigma}^2_k = \mathbf{b}_k^\top \mathbf{\Gamma}_g \mathbf{b}_k. \tag{18}$$

### S2. Technical details for "Correlation-adjusted cell-type–level association".

In Supplementary Note S1, we derived the marginal association statistic $Z_k$ for each cell type $k$ by fitting the linear model

$$\tilde{d}_i = \alpha_k + \tilde{y}_{ik}\gamma_k + \epsilon_{ik}, \qquad \epsilon_{ik} \sim N(0, \sigma^2_{\epsilon_k}), \tag{19}$$

and showed that, under the small-$R^2$ approximation, the summary-statistics-based Z-score can be written as

$$Z_k \approx \sum_l b_{kl} \frac{\hat{\sigma}_l}{\hat{\sigma}_k} \frac{\hat{\beta}_l}{\text{se}(\hat{\beta}_l)}. \tag{20}$$

The marginal model in Equation (19) models each cell type separately. However, predicted GReX across cell types are generally correlated. This dependence arises because (i) cell-type decomposition from bulk expression introduces shared uncertainty, (ii) cell-type-specific GReX are constructed from overlapping genetic predictors, and (iii) GReX prediction models across cell types are jointly trained affecting the performance of each other. Consequently, when we observe a signal in one cell type, we may similarly observe the signal in another cell type with the correlated marginal Z-scores $(Z_1, \ldots, Z_K)$. To account for this correlation structure, we consider a joint linear model that simultaneously includes predicted GReX from all cell types.

#### Joint linear model

Let $\tilde{\mathbf{d}} = (\tilde{d}_1, \ldots, \tilde{d}_n)^\top \in \mathbb{R}^n$ denote the phenotype vector in the GWAS cohort. Define the joint design matrix

$$\tilde{\mathbf{Y}} = \begin{pmatrix} 1 & \tilde{y}_{11} & \cdots & \tilde{y}_{1K} \\ \vdots & \vdots & \ddots & \vdots \\ 1 & \tilde{y}_{n1} & \cdots & \tilde{y}_{nK} \end{pmatrix} \in \mathbb{R}^{n\times(K+1)}, \tag{21}$$

where the first column corresponds to the intercept and the remaining $K$ columns correspond to predicted GReX for each cell type.

The joint regression model is

$$\tilde{\mathbf{d}} = \tilde{\mathbf{Y}}\boldsymbol{\eta} + \boldsymbol{\epsilon}, \qquad \boldsymbol{\epsilon} \sim N(\mathbf{0}, \sigma^2_{\text{Joint}}\mathbf{I}_n), \tag{22}$$

where $\boldsymbol{\eta} = (\eta_0, \eta_1, \ldots, \eta_K)^\top$ contains the intercept and cell-type–specific regression coefficients. Under ordinary least squares, the estimator is

$$\hat{\boldsymbol{\eta}} = (\tilde{\mathbf{Y}}^\top\tilde{\mathbf{Y}})^{-1}\tilde{\mathbf{Y}}^\top\tilde{\mathbf{d}}. \tag{23}$$

Define $\boldsymbol{\Sigma} = \tilde{\mathbf{Y}}^\top\tilde{\mathbf{Y}} \in \mathbb{R}^{(K+1)\times(K+1)}$. Then

$$\text{Var}(\hat{\boldsymbol{\eta}}) = \sigma^2_{\text{Joint}}\boldsymbol{\Sigma}^{-1}. \tag{24}$$

Let $S_{kk} = [\boldsymbol{\Sigma}^{-1}]_{kk}$ denote the $k$-th diagonal element of $\boldsymbol{\Sigma}^{-1}$. The joint Z-score for cell type $k$ is

$$\tilde{Z}_k = \frac{\hat{\eta}_k}{\text{se}(\hat{\eta}_k)}, \qquad \text{se}(\hat{\eta}_k) = \sigma_{\text{Joint}}\sqrt{S_{kk}}. \tag{25}$$

#### Linking marginal and joint statistics

From the marginal regression in Eq. (19), the OLS normal equation implies

$$\tilde{\mathbf{y}}_k^\top\tilde{\mathbf{d}} = \Omega_k\hat{\gamma}_k, \tag{26}$$

where $\Omega_k = \tilde{\mathbf{y}}_k^\top\tilde{\mathbf{y}}_k$. Stacking Eq. (26) across all cell types gives

$$\tilde{\mathbf{Y}}^\top\tilde{\mathbf{d}} = \text{diag}(\Omega_k)\hat{\boldsymbol{\gamma}}, \tag{27}$$

where $\hat{\boldsymbol{\gamma}} = (\hat{\gamma}_0, \ldots, \hat{\gamma}_K)^\top$. Substituting into Eq. (23), we obtain

$$\hat{\boldsymbol{\eta}} = \boldsymbol{\Sigma}^{-1} \operatorname{diag}(\Omega_k)\hat{\boldsymbol{\gamma}}. \tag{28}$$

Using the definition of the marginal Z-score,

$$\hat{\gamma}_k = Z_k \operatorname{se}(\hat{\gamma}_k) = Z_k \frac{\sigma_{\epsilon_k}}{\sqrt{\Omega_k}}, \tag{29}$$

we have

$$\operatorname{diag}(\Omega_k)\hat{\boldsymbol{\gamma}} = \operatorname{diag}(\Omega_k^{1/2}) \operatorname{diag}(\sigma_{\epsilon_k})\mathbf{Z}. \tag{30}$$

Therefore,

$$\hat{\boldsymbol{\eta}} = \boldsymbol{\Sigma}^{-1} \operatorname{diag}(\Omega_k^{1/2}) \operatorname{diag}(\sigma_{\epsilon_k})\mathbf{Z}. \tag{31}$$

#### Small-$R^2$ approximation

Under complex human diseases, each gene explains only a small proportion of phenotypic variance, implying $R_k^2$ and $R_{\text{Joint}}$ is small. As shown in S1, this implies

$$\frac{\sigma_{\epsilon_k}^2}{\sigma_{\text{Joint}}^2} = \frac{(1 - R_J^2)\sigma_d^2}{(1 - R_k^2)\sigma_d^2} = \frac{1 - R_J^2}{1 - R_k^2} \approx 1. \tag{32}$$

Substituting into Equation (31) gives

$$\hat{\boldsymbol{\eta}} \approx \sigma_{\text{Joint}} \boldsymbol{\Sigma}^{-1} \operatorname{diag}(\Omega_k^{1/2})\mathbf{Z}. \tag{33}$$

Dividing by $\operatorname{se}(\hat{\eta}_k) = \sigma_{\text{Joint}}\sqrt{S_{kk}}$ yields the joint Z-score approximation

$$\tilde{\mathbf{Z}} \approx \operatorname{diag}(S_{kk}^{-1/2})\boldsymbol{\Sigma}^{-1} \operatorname{diag}(\Omega_k^{1/2})\mathbf{Z}. \tag{34}$$

#### Ridge regularization

In practice, predicted GReX across cell types may be highly correlated, making $\boldsymbol{\Sigma}$ ill-conditioned and numerically unstable to invert. To stabilize estimation, we apply ridge regularization and define

$$\boldsymbol{\Sigma} = \tilde{\mathbf{Y}}^\top \tilde{\mathbf{Y}} + \lambda \mathbf{I}, \qquad \Omega_k = \tilde{\mathbf{y}}_k^\top \tilde{\mathbf{y}}_k + \lambda, \tag{35}$$

where $\lambda > 0$ is a small regularization parameter. All subsequent derivations follow analogously with the ridge-adjusted $\boldsymbol{\Sigma}$ and $\Omega_k$.

### S3. Technical details for "Inferring cell-type–specific association patterns."

To facilitate the interpretation of whether disease-gene associations are cell-type–specific or shared, we applied the probabilistic pattern-based framework. For $K$ cell types, we define $L = 2^K$ mutually exclusive binary association patterns,

$$\mathbf{q}_\ell = (q_{\ell 1}, \ldots, q_{\ell K}) \in \{0, 1\}^K, \quad \ell = 1, \ldots, L, \tag{36}$$

where $q_{\ell k} = 1$ indicates association in cell type $k$, and $q_{\ell k} = 0$ otherwise. For example, when $K = 2$, we have $L = 4$ and the four patterns are 00 (no association), 10 (cell type 1 only), 01 (cell type 2 only), and 11 (shared association). Each gene is assumed to arise from one and only one latent association pattern.

For gene $i$ in cell type $k$, let $p_{ik}$ denote the marginal association $P$-value. We transform

$$t_{ik} = -2\log(p_{ik}), \tag{37}$$

which follows a $\chi^2_2$ distribution under the null hypothesis and a mixture of non-central chi-squared distributions arising from heterogeneous genetic effects under the alternative distribution.

To understand the density function under the alternative distribution, we model the marginal distribution of $t_{ik}$ for each cell type $k$ using a two-component mixture. Specifically, letting $\theta_{1k}$ denote the genome-wide proportion of non-null associations in cell type $k$, the cumulative distribution function of $t_{ik}$ is modeled as

$$F_k(t; A_k, d'_k, \theta_{1k}) = (1-\theta_{1k})G(t;2) + \theta_{1k}G\left(\frac{t}{A_k}; d'_k\right), \tag{38}$$

where $G(\cdot;\nu)$ denotes the cumulative distribution function of a $\chi^2_\nu$ random variable. The null component is fixed as $\chi^2_2$, whereas the alternative distribution is approximated by a scaled chi-squared distribution $A_k\chi^2_{d'_k}$. The scaling factor $A_k > 0$ and degrees of freedom $d'_k$ are estimated via numerical optimization. To estimate $A_k\chi^2_{d'_k}$ and $d'_k$, we allow users to specify the nuisance parameter, $\theta_{1k}$, indicative of the small proportion of genes used to estimate the alternative. In our practice, we use half of the genes that are significance under 10% FDR. Additionally, our selection of the numerical optimization strategy focuses on minimizing discrepancies between empirical and model-based tail probabilities over the most extreme test statistics, ensuring robust approximation under sparse alternatives. This is due to that genome-wide associations are typically sparse (i.e., $\theta_{1k}$ is small), and accurate modeling of the extreme tail is more critical than matching lower-order moments.

With the estimated values, we are able to estimate the null and alternative density functions, $f_{0k}$ and $f_{1k}$, for each cell type $k$. Next, leveraging these estimated density functions for all $K$ cell types, based on transformed statistics for gene $i$, $\mathbf{T}_i = (t_{i1}, \ldots, t_{iK})^\top$, we derive a pattern-specific multivariate density $D_\ell(\mathbf{T}_i)$ for each patten $l \in (1, ..., L)$. When cell-type-specific density functions are independent,

$$D_\ell(\mathbf{T}_i) = \prod_{k=1}^{K} f_{0k}(t_{ik})^{1-q_{lk}}\, f_{1k}(t_{ik})^{q_{lk}},$$

indicating it's simply the product of the density function under each pattern. For example, when $K = 2$ and $L = 2^2 = 4$, $D_{00}(\mathbf{T}_i) = f_{01}(t_{ik})f_{02}(t_{ik})$ for pattern 00 (no association), $D_{10}(\mathbf{T}_i) = f_{11}(t_{ik})f_{02}(t_{ik})$ for pattern 10 (cell type 1 only), $D_{01}(\mathbf{T}_i) = f_{01}(t_{ik})f_{12}(t_{ik})$ for pattern 01 (cell type 2 only), $D_{11}(\mathbf{T}_i) = f_{11}(t_{ik})f_{12}(t_{ik})$ for pattern 11 (shared association). When cell-type-specific density functions are correlated, $D_\ell(\mathbf{T}_i)$ is extended using a multivariate gamma distribution, allowing dependence across cell types as in Primo.

Then, our goal is to use the estimated multivariate density functions to compute the probabilities that gene $i$ belongs to pattern $\ell$. We consider a Bayes' rule and the posterior probability for an association pattern $a_i \in \{1, \ldots, L\}$ for gene $i$ is

$$P(a_i = \ell \mid \mathbf{T}_i) = \frac{\pi_\ell D_\ell(\mathbf{T}_i)}{\sum_{b=1}^{L} \pi_b D_b(\mathbf{T}_i)}, \tag{39}$$

where $\pi_\ell$ is the prior transcriptome-wide proportion of genes belonging to pattern $\ell$, satisfying $\sum_{\ell=1}^{L} \pi_\ell = 1$. This mixing proportion $\pi_\ell$ captures the overall prevalence of different association configurations across the transcriptome. As both $\pi_\ell$ and the estimated pattern-specific densities $D_\ell(\mathbf{T}_i)$ are unknown, we use an Expectation–Maximization algorithm to iteratively estimate them. After convergency, the estimated pattern-specific probabilities provide a probabilistic interpretation of cell-type specificity for each gene.

#### Supplementary Figure 1

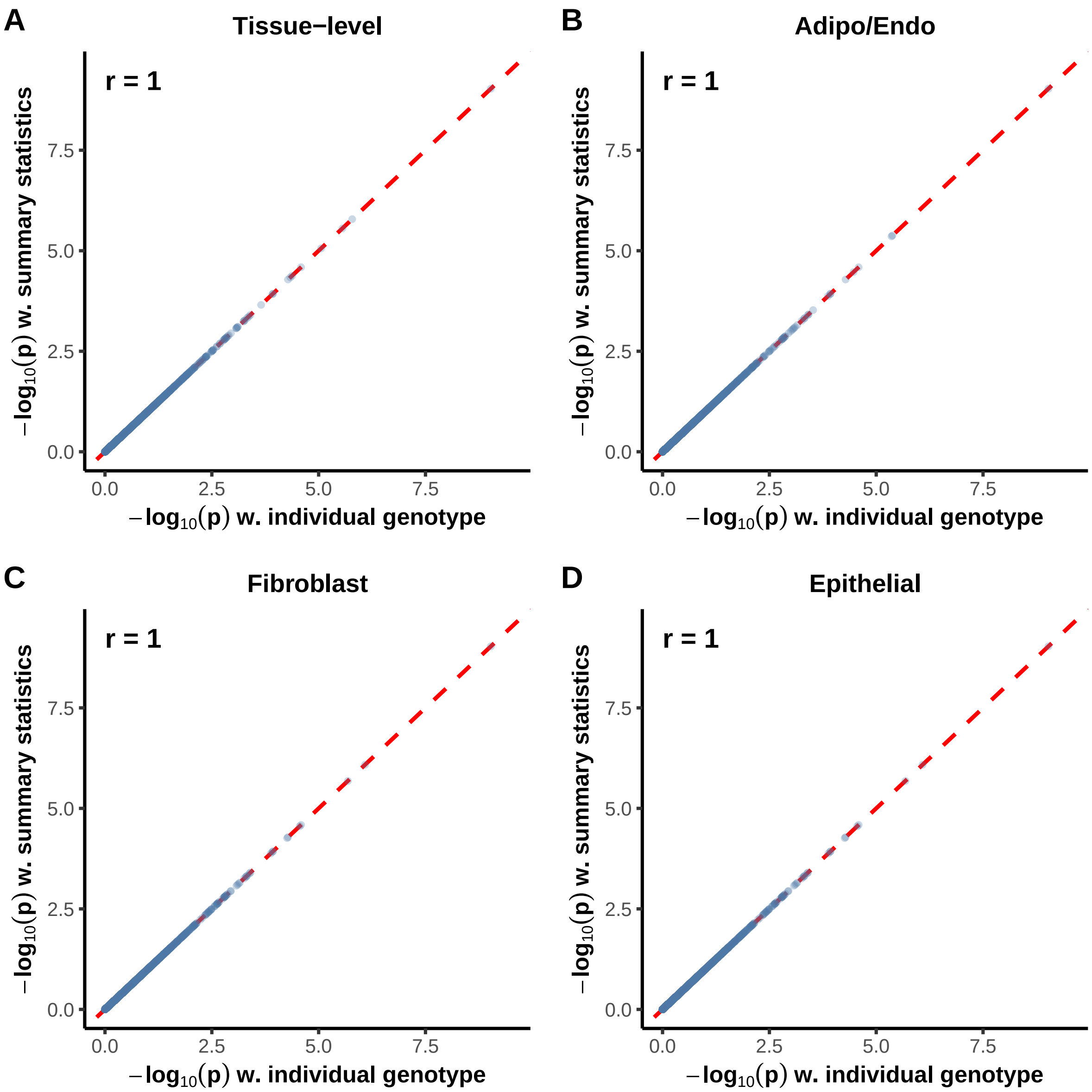


Figure S1: S-MiXcan that uses GWAS summary statistics produces p-values highly concordant with results derived from individual-level genotype data using MiXcan in the DRIVE cohort. (A) Scatter plot comparing $log_{10}(p)$ values from summary-statistics based inference in S-MiXcan and individual-level genotype based inference in MiXcan across all genes at the tissue level. Each point represents one gene, and the dashed red line indicates the line of equality (y = x). The Pearson correlation coefficient (r) quantifies concordance between the two methods. (B) Scatter plot comparing $log_{10}(p)$ values from summary-statistics based inference in S-MiXcan and individual-level genotype based inference in MiXcan across all genes in the adipocytes/endothelial cells. (C) Scatter plot comparing $log_{10}(p)$ values from summary-statistics based inference in S-MiXcan and individual-level genotype based inference in MiXcan across all genes in the fibroblast cells. (D) Scatter plot comparing $log_{10}(p)$ values from summary-statistics based inference in S-MiXcan and individual-level genotype based inference in MiXcan across all genes in the epithelial cells.

### Supplementary Figure 2

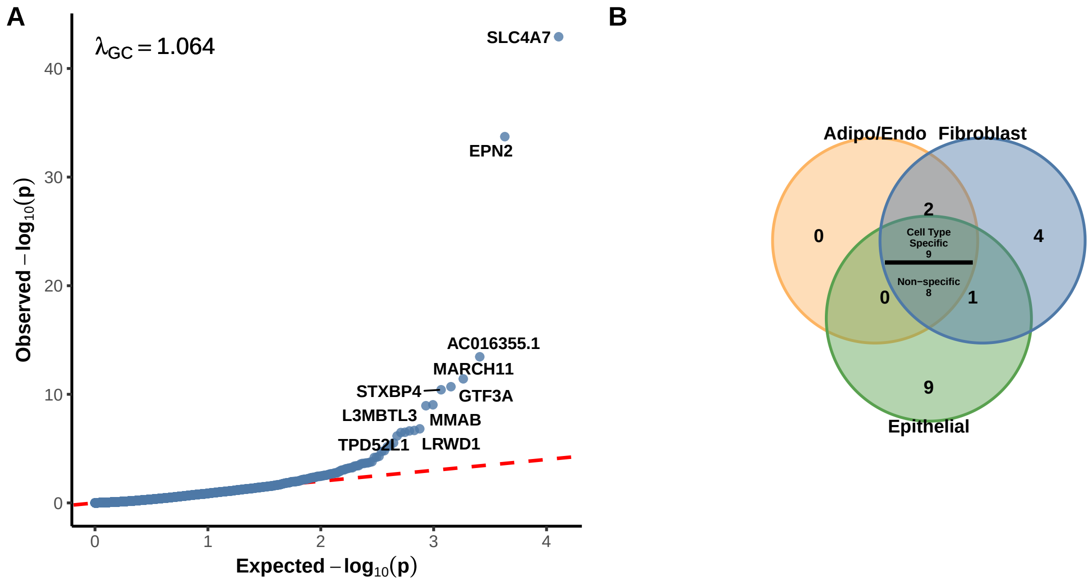


Figure S2: Performance of S-MiXcan under three cell types with BCAC meta-analysis GWAS summary statistics. (A) Application of S-MiXcan to BCAC meta-analysis GWAS summary statistics maintains well-controlled type I error rates ($\lambda_{GC} = 1.063$) with significant association signals. (B) Inferred cell-type-specificity among 33 suggestive genes at 10% FDR.
